# Early postmortem brain MRI findings in COVID-19 non-survivors

**DOI:** 10.1101/2020.05.04.20090316

**Authors:** Tim Coolen, Valentina Lolli, Niloufar Sadeghi, Antonin Rovaï, Nicola Trotta, Fabio Silvio Taccone, Jacques Creteur, Sophie Henrard, Jean-Christophe Goffard, Olivier De Witte, Gilles Naeije, Serge Goldman, Xavier De Tiège

## Abstract

**Importance:** The severe acute respiratory syndrome coronavirus 2 (SARS-CoV-2) is considered to have potential neuro-invasiveness that might lead to acute brain disorders or contribute to respiratory distress in patients with coronavirus disease 2019 (COVID-19). Brain magnetic resonance imaging (MRI) data in COVID-19 patients are scarce due to difficulties to obtain such examination in infected unstable patients during the COVID-19 outbreak.

**Objective:** To investigate the occurrence of structural brain abnormalities in non-survivors of COVID-19 in a virtopsy framework.

**Design:** Prospective, case series study with postmortem brain MRI obtained early (<24h) after death.

**Setting:** Monocentric study.

**Participants:** From 31/03/2020 to 24/04/2020, consecutive decedents who fulfilled the following inclusion criteria were included: death <24 hours, SARS-CoV-2 detection on nasopharyngeal swab specimen, chest computerized tomographic (CT) scan suggestive of COVID-19, absence of known focal brain lesion, and MRI compatibility.

**Main Outcome(s) and Measure(s):** Signs of acute brain injury and MRI signal abnormalities along the olfactory tract and brainstem were searched independently by 3 neuroradiologists, then reviewed with neurologists and clinicians.

**Results:** Among the 62 patients who died from COVID-19 during the inclusion period, 19 decedents fulfilled inclusion criteria. Subcortical micro- and macro-bleeds (2 decedents), cortico-subcortical edematous changes evocative of posterior reversible encephalopathy syndrome (PRES, one decedent), and nonspecific deep white matter changes (one decedent) were observed. Asymmetric olfactory bulbs were found in 4 other decedents without downstream olfactory tract abnormalities. No brainstem MRI signal abnormality.

**Conclusions and Relevance:** Postmortem brain MRI demonstrates hemorrhagic and PRES-related brain lesions in non-survivors of COVID-19 that might be triggered by the virus-induced endothelial disturbances. SARS-CoV-2-related olfactory impairment seems to be limited to olfactory bulbs. The absence of brainstem MRI abnormalities does not support a brain-related contribution to respiratory distress in COVID-19.

**Key Points:** *Question:* Is there common brain MRI abnormalities patterns in non-survivors of coronavirus disease 2019 ?

*Findings:* In a case series of 19 non-survivors of severe COVID-19 disease, early postmortem brain MRI demonstrated patterns evocative of intracranial vasculopathy in 4 decedents: subcortical micro- and macro-bleeds (2 decedents), cortico-subcortical edematous changes evocative of posterior reversible encephalopathy syndrome (PRES, one decedent), and nonspecific deep white matter changes (one decedent). Asymmetric olfactory bulbs were found in 4 other decedents but without downstream olfactory tract abnormalities.

*Meaning:* Postmortem brain MRI demonstrates hemorrhagic and PRES-related brain lesions in non-survivors of COVID-19 that might be triggered by virus-induced endothelial disturbances.

## Introduction

Infection by the severe acute respiratory syndrome coronavirus 2 (SARS-CoV-2), named coronavirus disease 19 (COVID-19) by the World Health Organization, is associated with an ongoing worldwide outbreak of atypical and severe pneumonia.^1^

SARS-CoV-2 uses human cell receptor angiotensin-converting enzyme 2 (ACE2) as cellular entry point.^1^ ACE2 is expressed in airway epithelia, lung parenchyma, vascular endothelium, heart, kidneys, and small intestine^2^, which explains most of COVID-19 symptoms. ACE2 is also present in glial cells and neurons of mammalian brains, especially in brainstem nuclei involved in cardio-respiratory function.^2^ SARS-CoV-2 is therefore considered to have potential neuro-invasiveness that might lead to acute brain disorders or contribute to respiratory distress in COVID-19 patients.^3^ SARS-CoV-2 might enter the central nervous system (CNS) through hematogenous dissemination or neuronal retrograde route via, e.g., olfactory bulbs or medullary neurons.^3^ The anosmia frequently observed in SARS-CoV-2 infected patients^4^ supports the neural retrograde hypothesis. The neuro-invasiveness of SARS-CoV-2 is also supported by case reports of meningo-encephalitis,^5^ intracerebral hemorrhage,^6^ and secondary acute necrotizing encephalopathy^7^ associated with SARS-CoV-2 infection. Central neurological symptoms (e.g., headache, stroke, impaired consciousness) are also observed in 25% of COVID-19 patients.^8^ Still, brain magnetic resonance imaging (MRI) data are scarce in COVID-19 patients due to difficulties to obtain such examination in infected unstable patients during the COVID-19 outbreak.^8^

This prospective, monocentric, postmortem brain MRI case series investigated the occurrence of structural brain abnormalities in non-survivors of COVID-19 in a brain virtopsy framework performed early (≤24h) after death. We specifically searched for signs of acute brain injury (e.g., stroke, encephalitis) and MRI signal abnormalities along the olfactory tract and brainstem.

## Methods

### Study Design and participants

This study was carried out at the CUB Hôpital Erasme (Brussels, Belgium) after approval by the institutional Ethical Committee (Ref: P2020/204, SRB2020121), which did not request informed consent from legal representatives.

From 31/03/2020 to 24/04/2020, consecutive decedents who fulfilled the following inclusion criteria were included: death <24 hours; SARS-CoV-2 detection (direct antigen detection or reverse transcriptase polymerase chain reaction) on nasopharyngeal swab specimen, chest computerized tomographic (CT) scan suggestive of COVID-19 without alternative diagnosis; absence of known focal brain lesion; MRI compatibility. Clinical data were retrieved from decedents’ medical record and reviewed by clinicians (F.S.T., S.H., J.C.G., G.N., O.D.). See eMethods for clinical definitions.

After death, decedents were directly placed in an MRI-compatible mortuary bag, brought to the morgue, and placed in dedicated mortuary refrigerators (2-3 C°).

### MRI data acquisition

Postmortem structural brain MRI were performed on a 3 Tesla hybrid PET-MR scanner (SIGNA^TM^, GE Healthcare) in accordance with COVID-19 institutional hygienic and safety rules. The PET-MR facility was dedicated to research investigations in COVID-19 patients. Mortuary bags were not opened during the procedure.

Whole-brain (high-resolution 3D T1-weighted imaging (T1WI), axial T2-weighted imaging (T2WI), sagittal 3D T2WI fluid-attenuated inversion recovery (FLAIR), axial 3D susceptibility-weighted imaging (SWI), axial diffusion-weighted imaging (DWI)) and olfactory bulb-focused (coronal T2WI) MRI sequences were acquired in all decedents (eMethods for details).

### MRI data analyses

MRI data were first independently reviewed by three neuroradiologists (V.L., T.C., N.S.) based on systematic and comprehensive visual assessment (eMethods). Final reports were then discussed with three neurologists (S.G., X.D.T., G.N.) and with clinicians (F.S.T., S.H., J.C.G., O.D.).

## Results

Among the 62 patients who died from COVID-19 infection during the inclusion period, 19 decedents fulfilled inclusion criteria (eResults). They spent from 2h04 to 23h45 hours in mortuary refrigerators before neuroimaging.

Table 1 summarizes decedents’ characteristics (eResults and eTable 1 for details).

All decedents had severe COVID-19 with 1 up to 5 comorbidities (i.e., hypertension, heart disorder, diabetes, chronic obstructive pulmonary disorder, chronic kidney disease, or body mass index >25). Decedents developed COVID-19 complications (coagulopathy (6/19), acute kidney injury (14/19) or acute cardiac injury (18/19)). Fifteen decedents died from respiratory failure, while 4 others died from septic shock and multiorgan failure.

MRI abnormalities related to postmortem status are described in eResults, eFigures 1-4 and eTable 2. Figure 1 illustrates structural cerebral abnormalities found in COVID-19 non-survivors (see also eResults, eFigures 5-6, eTable 3).

**Figure 1:**
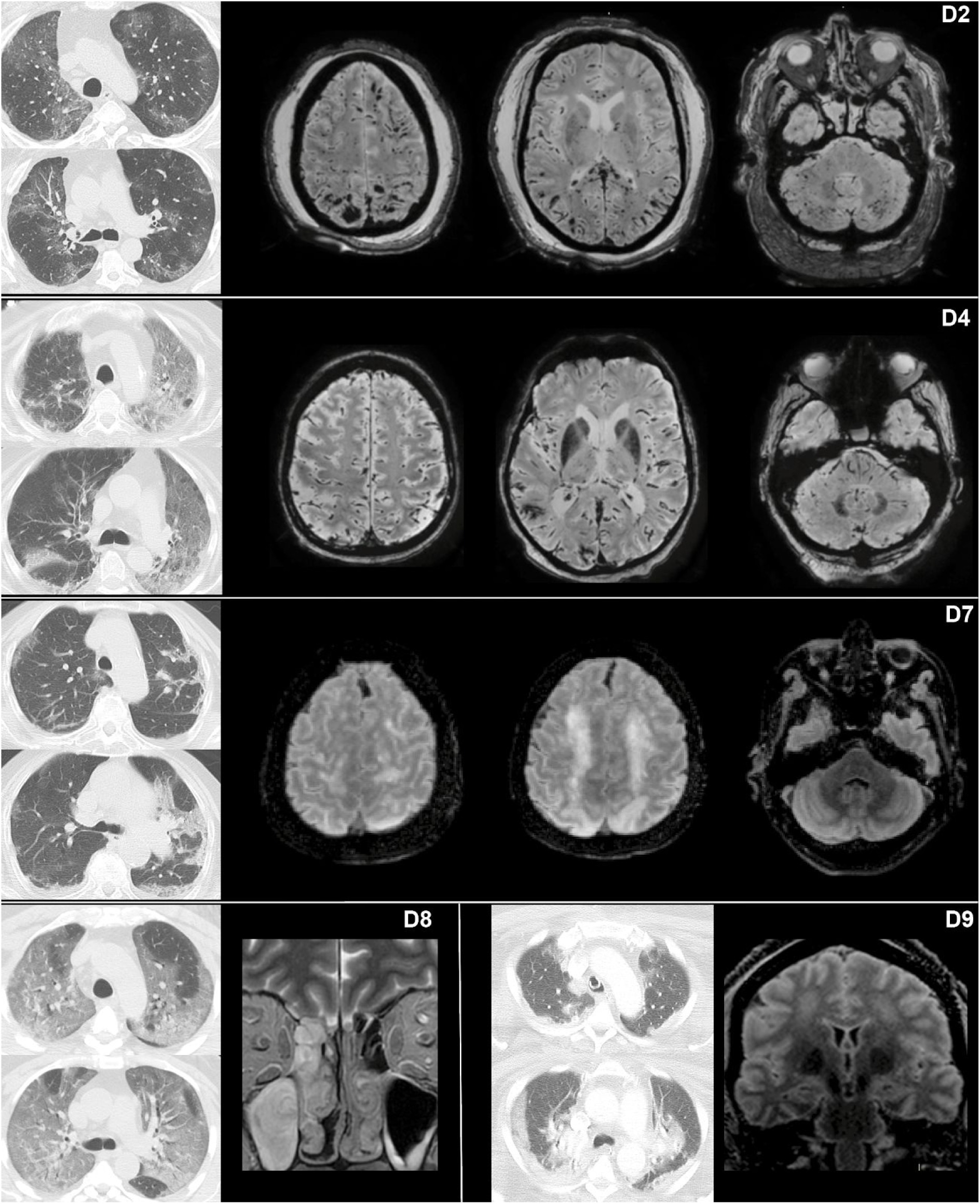
Illustration of postmortem brain MRI findings in five decedents (D2, D4, D7, D8, D9). For each decedent, two axial slices of their chest CT scan (first two images on the left) are provided to illustrate the typical SARS-CoV-2-related ground-glass multifocal lung opacities (delay between chest CT scan and death: 10 days, D2; 1 day D4; 7 days, D7; 21 days, D8; 2 days, D9). D2 (first line) had diffuse micro- and macro-bleeds with parieto-occipito-temporal predominance, also involving the splenium of corpus callosum, left superior frontal sulcus and bilateral cerebellum, as demonstrated on the susceptibility weighted imaging (SWI) sequence. D4 (second line) presented with two isolated arciform subcortical macro-bleeds in the right occipital and temporal lobes, also visible on the SWI sequence. D7 (third line) displayed cortical and subcortical high FLAIR signal intensity and swelling in the bilateral superior precentral gyri and superior parietal lobules, while the posterior fossa was spared. D8 (fourth line, left) had a relatively inflated left olfactory bulb associated with ipsilateral obliteration of the olfactory cleft. D9 (fourth line, right) showed bilateral, extensive and hazy FLAIR high signal intensity of the bilateral centrum semiovale. All images are displayed in radiological convention.

Parenchymal brain MRI abnormalities were found in four decedents. Decedent 2 (D2) had diffuse micro- and macrobleeds with subcortical and posterior predominance. D4 presented two subcortical arciform bleeds in the right occipito-temporal region. D7 showed superior precentral and parietal cortico-subcortical swelling associated with marked supratentorial white matter changes. D9 had extensive T2WI hazy hyperintensity in the bilateral centrum semiovale.

Four decedents had asymmetric olfactory bulbs with ipsilateral (D8) or without (D5, D11, D12) olfactory cleft obliteration. No other abnormality was found along the olfactory tract.

No signal abnormality was found in the brainstem, except in D10 who had a midline pontine signal abnormality evocative of capillary telangiectasia (eFigure 6).

## Discussion

Parenchymal brain MRI abnormalities evocative of intracranial vasculopathy were observed in 4/19 decedents (21%). Asymmetric olfactory bulbs were found in 4 others (21%).

D2 had diffuse subcortical micro- and macro-bleeds with posterior predominance. Considering his clinical context, these abnormalities were evocative of multifocal hemorrhagic lesions triggered by disseminated intravascular coagulation (DIC),^9,10^ eventually favored by veno-venous extracorporeal membrane oxygenation.^11^ DIC is indeed frequently observed in severe COVID-19.^10^

In D4, we observed two posterior subcortical bleeds — unlikely due to small vessel disease or amyloid angiopathy —, which shared some similarities with those observed in D2.

D7 showed superior precentral and parietal cortico-subcortical swelling associated with marked supratentorial white matter changes. These findings are evocative of posterior reversible encephalopathy syndrome (PRES). PRES refers to reversible vasogenic brain oedema predominating in posterior brain regions, which occurs in association with a variety of systemic disorders.^12^ PRES is typically caused by endothelial injury related to abrupt blood pressure changes or direct effects of cytokines on the endothelium.^12^

D9 had extensive hazy T2WI hyperintensity of the centrum semiovale, probably reflecting early edematous changes or deep watershed infarction.

In those 4 decedents, SARS-CoV-2-induced intracranial vasculopathy is the suspected common denominator between DIC-related lesions, PRES and centrum semiovale hyperintensity. The widespread expression of ACE2 in endothelial cells may be the trigger of virus-induced endothelial dysfunction in COVID-19.^13^ So, the vasculopathy could be induced by direct SARS-CoV-2 infection of endothelial cells, with secondary inflammation and activation, or by a systemic cytokine storm. Both occur in severe COVID-19 leading to a widespread “endotheliitis”.^13^

Asymmetric olfactory bulbs with or without olfactory cleft obliteration were observed in 4/19 decedents. This may represent an MRI-correlate of the anosmia/hyposmia frequently observed in SARS-CoV-2 patients and may support the neural retrograde hypothesis for SARS-CoV-2 CNS dissemination. Still, this study did not find any MRI signal abnormality downstream the olfactory tract. It also failed to find specific brainstem abnormalities, which does not support a brain-related contribution to respiratory distress in COVID-19.^3^

Further discussion and study limitations are available in eDiscussion.

In summary, this postmortem MRI study demonstrated the occurrence of vasculopathic brain lesions in COVID-19 non-survivors that might be triggered by the endothelial disturbances associated with SARS-CoV-2 infection.

## Data Availability

Data are available upon reasonable request and if accepted by the Ethical Committee

## Acknowledgments

The authors would like to warmly thank all the personnel of the morgue and of the PET-MR Unit of the CUB Hôpital Erasme. They have done a fantastic job to make this study possible.

## Author Contributions

Tim Coolen, Valentina Lolli, Niloufar Sadeghi, Serge Goldman and Xavier De Tiège had full access to all of the data in the study and take responsibility for the integrity of the data and the accuracy of the data analysis.

Tim Coolen and Valentina Lolli contributed equally and share first authorship.

### Concept and design

Tim Coolen, Valentina Lolli, Niloufar Sadeghi, Olivier De Witte, Serge Goldman and Xavier De Tiège

### Acquisition and analysis

Tim Coolen, Valentina Lolli, Niloufar Sadeghi, Serge Goldman and Xavier De Tiège

### Interpretations of data

all authors

### Drafting the manuscript

Tim Coolen, Valentina Lolli, Niloufar Sadeghi, Serge Goldman and Xavier De Tiège

### Critical revision of the manuscript for important intellectual content

all authors.

### Statistical analyses

Tim Coolen

### Obtained funding

Serge Goldman and Xavier De Tiège

### Supervision

Niloufar Sadeghi, Serge Goldman and Xavier De Tiège

